# Comparative analysis of the spatial distribution of brain metastases across several primary cancers using machine learning and deep learning models

**DOI:** 10.1101/2023.09.19.23295748

**Authors:** Saeedeh Mahmoodifar, Dhiraj J. Pangal, Jeremy Mason, Bodour Salhia, Josh Neman, Gabriel Zada, Paul K. Newton

## Abstract

**Objective:** Brain metastases (BM) are associated with poor prognosis and increased mortality rates, making them a significant clinical challenge. Therefore, studying BMs can aid in developing better diagnostic tools for their early detection and monitoring. Systematic comparisons of anatomical distributions of BM from different primary cancers, however, remain largely unavailable.

**Methods:** To test the hypothesis that anatomical BM distributions differ based on primary cancer type, we analyze the spatial coordinates of BMs for five different primary cancer types along principal component (PC) axes which optimizes their largest spread along each of the three PC axes. Data used in this analysis is taken from the International Radiosurgery Research Foundation (IRRF) and all patients underwent gamma-knife radiosurgery (GKRS) for the treatment of BMs which are labeled based on the primary cancer types Breast, Lung, Melanoma, Renal, and Colon. The dataset consists of six features including sex, age, target volume, and stereotactic Cartesian coordinates X, Y, and Z of a total of 3949 intracranial metastases. We employ PC coordinates to reduce the dimensionality of our dataset and highlight the distinctions in the anatomical spread of BMs between various cancer types. We utilized different Machine Learning (ML) algorithms: Random Forest (RF), Support Vector Machine (SVM), and TabNet Deep Learning (DL) model to establish the relationship between primary cancer diagnosis, spatial coordinates of BMs, age, and target volume.

**Results:** Our findings demonstrate that the first principal component (PC1) exhibits a greater alignment with the Y axis, followed by the Z axis, with a minimal correlation observed with the X axis. Based on our analysis of the PC1 versus PC2 plots, we have determined that the pairs of Breast and Lung cancer, as well as Breast and Renal cancer, exhibit the most notable distinctions in their anatomical spreading patterns. In contrast, we find that the pairs of Renal and Lung cancer, as well as Lung and Melanoma, were most similar in their patterns. Our ML and DL results indicate high accuracy in distinguishing the distribution of BM for different primary cancers, with the SVM algorithm achieving a 97% accuracy rate when using a polynomial kernel and TabNet a 96% accuracy. The RF algorithm ranks PC1 as the most important discriminating feature.

**Conclusions:** Taken together, the results demonstrate an accurate multiclass machine learning classification with respect to the distribution of brain metastases.

## I. INTRODUCTION

It is well established that different primary cancer types, and different molecular subtypes distribute metastases preferentially to different locations [1–8], although a quantitative understanding of the spatial distribution of metastatic disease, and the temporal ordering of when these metastases first appear at the different locations remains far less understood [3]. Although metastases to the brain are not usually the location of the first metastatic site for any primary cancer type [4], the presence of BMs portend poor prognosis for the patient, regardless of cancer subtype. The advancements in treatment regimens, including the development of immunologic therapies have increased life expectancies for a number of primary cancers, and brought new importance to the study of BMs, their natural progression and causes for growth.

Recently, progress has been made in quantifying the spatial distribution of brain metastases for breast cancer patients of different molecular subtypes [9], showing quantitively distinct patterns in some categories. The underlying hypothesis rests on the notion that different cancers require different environments for growth [10, 11], and therefore are more or less likely to metastasize in certain regions of the brain. We aim to expand on the work performed by our group in prior studies [1–9], by exploring the predictive ability of a machine learning model to determine the primary subtype of cancer given spatial information about its three-dimensional location, as well as age at treatment and target volume. The potential ability for machine learning models to accurately identify the primary cancer type from these small set of features would indicate that these differences are distinct enough to be discerned, which might further motivate the search for underlying biological explanations for these differences.

We demonstrate that using spatial data as the primary means of input, a machine learning model can accurately parse out the primary cancer subtype from a large dataset of brain metastases from a national brain tumor metastasis registry.

## II. METHODS

### A. Dataset

Data used in this analysis is taken from IRRF and all patients underwent GKRS for the treatment of brain metastases which are labeled based on the primary cancer types Breast, Lung, Melanoma, Renal, and Colon. The dataset consists of six features including sex, age, target volume, and stereotactic Cartesian coordinates X, Y, and Z of a total of 3949 intracranial metastases. See the data summarized in Table I and Table II.

**TABLE I.** Number of brain metastases and proportion of sex subgroupings for different primary cancer types.

| Diagnosis | Sex | Total number of mets |
| --- | --- | --- |
| Breast | Female | 550 |
|  | Male | 0 |
| Colon | Female | 56 |
|  | Male | 56 |
| Lung | Female | 478 |
|  | Male | 588 |
| Melanoma | Female | 646 |
|  | Male | 1242 |
| Renal | Female | 66 |
|  | Male | 267 |

**TABLE II.** Characteristics of the study population.

| | X (mm) | Y (mm) | Z (mm) | PC 1 | PC 2 | PC 3 | Age at Treatment (yrs) | Target volume ($cm^3$) |
| --- | --- | --- | --- | --- | --- | --- | --- | --- |
| Total number of mets | 3949 | 3949 | 3949 | 3949 | 3949 | 3949 | 3949 | 3949 |
| mean | 99.39 | 88.94 | 112.96 | 0.00 | 0.00 | 0.00 | 57.57 | 3.44 |
| median | 99.50 | 85.30 | 112.90 | 0.12 | -0.65 | -0.47 | 58.00 | 0.61 |
| std | 28.39 | 36.09 | 31.27 | 37.85 | 29.18 | 27.76 | 13.07 | 13.13 |
| min | 32.70 | 4.50 | 20.60 | -95.40 | -72.59 | -72.66 | 13.00 | 0.00 |
| max | 188.80 | 190.10 | 199.70 | 102.03 | 101.71 | 81.79 | 90.00 | 357.90 |

### B. Principal component analysis (PCA)

The principal component coordinates are a data driven orthogonal coordinate system intended to highlight the directions of the greatest spread of the data, with PC1 as the direction of the largest variance and PC2 and PC3 as the directions that capture the remaining variations orthogonal to the first principal component and to each other. PCA is used to identify patterns in a dataset and as a method of dimensionality reduction for high dimensional datasets by identifying new uncorrelated features (PC), which allows better visualization of the dataset [12]. We use PCA from the Scikit-learn library in Python for our analysis [13].

### C. Synthetic Minority Over-sampling Technique (SMOTE)

In essence, SMOTE is a data augmentation method used to address a class imbalance in supervised machine learning problems. Class imbalance occurs when one class of a classification problem has significantly fewer samples than the other classes, which can lead to poor performance of the classifier on the minority class. SMOTE creates synthetic samples of the minority class by interpolating between existing minority class samples. The method selects a minority class sample and identifies its k nearest neighbors in the feature space. SMOTE then creates a new sample by randomly selecting one of the “k” nearest neighbors and creating a synthetic sample between the original sample and each of its neighbors that is a linear combination of the original and selected neighbors. The process repeats until the desired balance between the classes is achieved. The synthetic samples created by SMOTE increase the size of the minority class, making it more representative and improving the classifier’s ability to learn the patterns in the minority class [14].

### D. Dataset Preprocessing

PCA is sensitive to the scaling of the variables in the dataset. Variables that have larger magnitudes will dominate the variance and may obscure the contribution of other variables that have smaller magnitudes. Scaling the variables to a common scale ensures that all variables are equally important in the analysis. Different variables in the dataset have different units of measurement, and these units can affect the calculation of the principal components. Scaling the variables to unit variance (i.e., standardizing) removes the units of measurement and allows the components to be calculated based on the correlations between the variables. In order to ensure that the result of the PCA is representative of the underlying patterns in the data, we scale our features before performing the PCA. We use the StandardScaler from the Scikitlearn package in Python which set the mean to zero and the variance to one [13]. In order to reduce the effect of class imbalance in the dataset, we use the Synthetic Minority Over-sampling Technique using the SMOTE from the imbalanced-learn library in Python [15]. We split up the dataset into 90% training and 10% testing. The reported evaluation metrics in Table III correspond to the testing dataset.

**TABLE III.** Evaluation metrics for different machine learning and deep learning models.

| Model | Accuracy | Precision | Recall | F1-score |
| --- | --- | --- | --- | --- |
| RF | 89% | 89% | 89% | 89% |
| SVM-linear | 71% | 78% | 71% | 72% |
| SVM-poly | 97% | 97% | 97% | 97% |
| SVM-rbf | 94% | 94% | 94% | 94% |
| TabNet | 96% | 97% | 96% | 96% |

### E. Random Forest (RF)

Random Forest is a supervised machine learning algorithm for classification and regression tasks. It is an ensemble learning method that combines multiple decision trees to make a final prediction. During the training process, Random Forest builds a large number of decision trees by using a randomly selected portion of the training data along with a randomly selected subset of the available features. Each tree is built independently, and at each split, the algorithm selects the best feature to split on among a random subset of features. This randomness helps reduce overfitting and improves the model’s generalization performance. Once the trees are built, the Random Forest algorithm combines their predictions to make a final prediction. In classification tasks, the class with the most votes is selected, and in regression tasks, the mean or median of the individual tree predictions is taken [16]. A random forest classifier is built based on RandomForestClassifier from the Scikit-learn package [13].

### F. Support Vector Machine (SVM) and One v. All (OvA)

The linear SVM algorithm aims to find a hyperplane that separates two tumor classes to maximize the distance between the hyperplane and the nearest samples from each class. In order to determine the maximum separation distance between classes, the dot products of support vectors and the classes must be computed [17]. The main concept behind this is identifying the largest margin between the classes. In cases where the data is not linearly separable, SVMs can use a kernel function to transform the data into a higher dimensional space that a hyperplane can separate. The kernel functions used in this study are linear, polynomial, and radial basis functions. For transitioning from binary to multiclass classification, we adopt a One-vs-All (OvA) approach [18]. The OvA strategy involves training multiple binary classifiers, each distinguishing one class from all the others. For each class, a binary classifier is trained to distinguish between that class and all the other classes combined. This results in a set of binary classifiers, one for each class. The classifier with the highest confidence score is selected as the predicted class during prediction. This study uses three different kernels: Radial Basis Function, Polynomial, and Linear kernel. For our analysis, we utilize the SVC algorithm from the Scikit-learn library in Python [13].

### G. TabNet

Deep learning algorithms have generally been successful in classifying images or audio but not tabular data [19]. TabNet, a deep neural network (DNN) tailored for learning from tabular data, employs a distinctive architecture known as the TabNet encoder [20]. In this architecture, sequential multi-steps (Nsteps) are a pivotal component. Each step, denoted as *i*, leverages processed information from the previous step (*i −*1) to make decisions regarding feature utilization. These decisions culminate in processed feature representations, which, in turn, play a critical role in the overarching decision-making process. Notably, the model ingests a dataset characterized by a specific batch size (B) and D-dimensional features as input without applying global feature normalization. Subsequently, the data undergoes batch normalization (BN) before being channeled into a feature transformer. Within the feature transformer, illustrated in Figure 4c of the original paper [20], several gated linear unit (GLU) blocks are employed (a total of n). Each GLU block encompasses three essential layers: fully connected (FC), batch normalization (BN), and GLU. In cases where four GLU blocks are used, two are shared, and two operate independently, contributing to the model’s robustness and parameter-efficient learning. A skip connection is also established 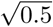 between consecutive blocks. Following each block, a 0.5 normalization process ensues, ensuring stability by maintaining variance. Subsequently, the feature transformer processes the batch-normalized features and transfers this information to the attentive transformer of the current step through a split layer. The attentive transformer, visualized in Figure 4d of the original paper [20], comprises four layers: FC, BN, prior scales, and sparsemax. It accepts input from the split layer, applies FC and BN layers, and then employs the prior scales layer to consolidate the magnitudes of features utilized in preceding decision steps. This consolidation is governed by the equation 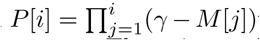 where *γ* denotes a relaxation parameter. The primary function of the attentive transformer is to compute the mask layer for the current step, utilizing the outcomes from the previous step. The learnable mask (*M* [*i*]) is instrumental for sparse selection of the most pertinent features, thereby enhancing parameter efficiency by prioritizing relevant features. The multiplicative masking process involves the attentive transformer to derive masks using processed features from the prior step (*a*[*i* 1]). This process is detailed in the equation *M* [*i*] = *sparsemax*(*P* [*i −* 1].*h*_*i*_(*a*[*i* − 1])) where *P* [*i*] is the priori scale and hi some trainable function. These masks contribute to model interpretability, with TabNet offering both local and global explanations. Local interpretability is achieved by utilizing TabNet’s decision masks. TabNet is designed to provide a comprehensive framework for feature selection and efficient learning from tabular data, and its unique encoder architecture, featuring sequential multi-steps and attentive transformers, contributes to its effectiveness in these tasks.

## III. RESULTS

In figure 1 we show 2D scatter plots of BM locations for five different primary cancer types (Breast, Lung, Melanoma, Renal, and Colon) plotted with their PC1 component versus each of the (X, Y, Z) Cartesian coordinates in 3D space. Figure 1(a) shows that PC1 correlates most strongly with the Y coordinate (front-to-back), next, figure 1(b) shows the correlation with the Z coordinate (top-to-bottom), while figure 1(c) shows there is very little linear correlation with the X coordinate (side-to-side). See [9] Figure S1 for a more detailed plot of the coordinate systems used with the Gamma Knife radiosurgery (GKRS) stereotactic headset which measures BM locations. Our conclusion from these comparisons is that the (PC1, PC2) plane offers an optimal [12] reduced dimension plane that most accurately will depict the differences in the spatial distributions of BMs for the five different cancer types, in addition to the other features from the data set. The side-to-side X coordinate distribution is the least important of the three, reflecting the fact that the five cancers all distribute their BMs more or less symmetrically across the midline. Given that it is the (Y, Z) coordinate plane that mostly captures the important differences in Cartesian BM locations, we show in figure 2 PC1 for each of the primary cancer types projected onto this plane. Both the means (the base of each coordinate arrow) and the directions of each PC1 vector are different for each primary cancer type as can easily be seen.

**FIG. 1.**
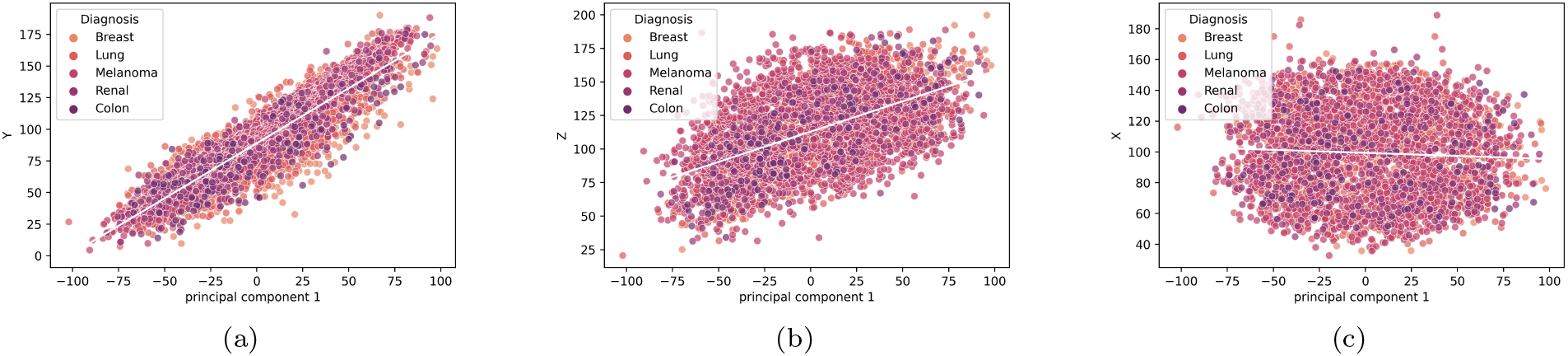
Brain metastases scatter plots for five different primary cancers along Principal Component 1 axis (PC1) vs. X, Y, and Z coordinates showing strongest linear correlation between Y axis and PC1 axis. a) 2D projection of data onto (PC1, Y) plane and linear curve fit; b) 2D projection of data onto (PC1, Z) plane and linear curve fit; c) 2D projection of data onto (PC1, X) and linear curve fit.

**FIG. 2.**
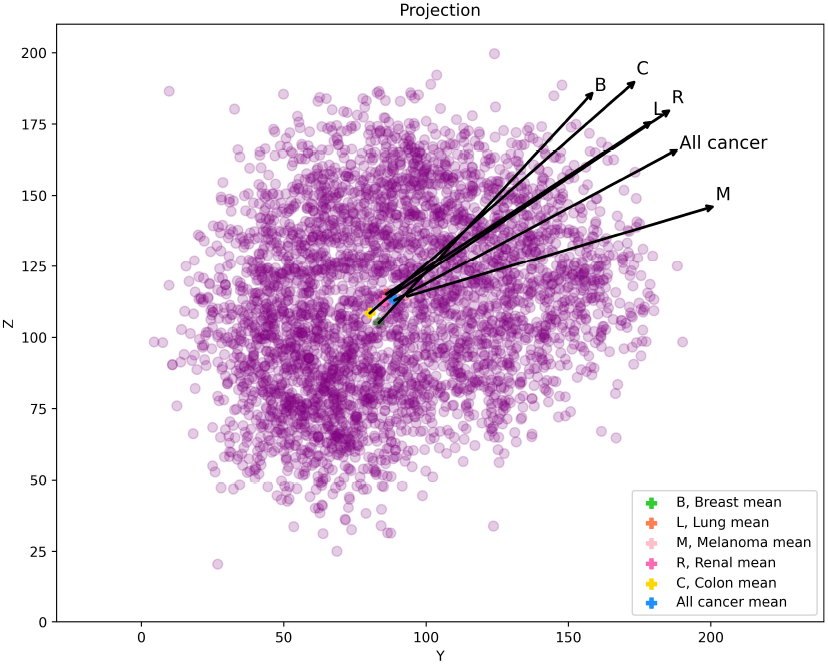
2D projection of scatter plot of all cancer metastatic brain tumors onto (Y, Z) plane showing the Principal component 1 axis for each cancer type separately and with respect to all cancer types together. Violet crosses indicate the means of each cancer and the yellow cross indicates the mean of all data points.

Note the similarity, however, between the direction of the PC1 axis associated with lung and renal cancers, with only the mean basepoint shifting between the two.

In figure 3 we focus on depicting the BM locations in the PC1 vs. PC2 planes (the optimal reduced-order plane). In figures 3(a), 3(b) we show the distributions of the two cancer types that are most distinct with respect to their spatial distributions: Breast vs. Lung (3(a)), and Breast vs. Renal (3(b)). We indicate these differences by plotting the linear curve fits to each of the cancer types on the same plots, showing both their means and orientation of the linear curves are very distinct. By contrast, figures 3(c), 3(d) show the distributions that are most similar: Lung vs. Melanoma (3(c)), and Lung vs. Renal (3(d)). Note the similarity of their means and linear curve fits as compared with those in figures 3(a), 3(b). The linear curve fits in these four plots are not intended to indicate that the data closely follows a linear regression model, but only meant to show the most apparent differences/similarities in the spread of points along the regression line (i.e a useful visual guide).

**FIG. 3.**
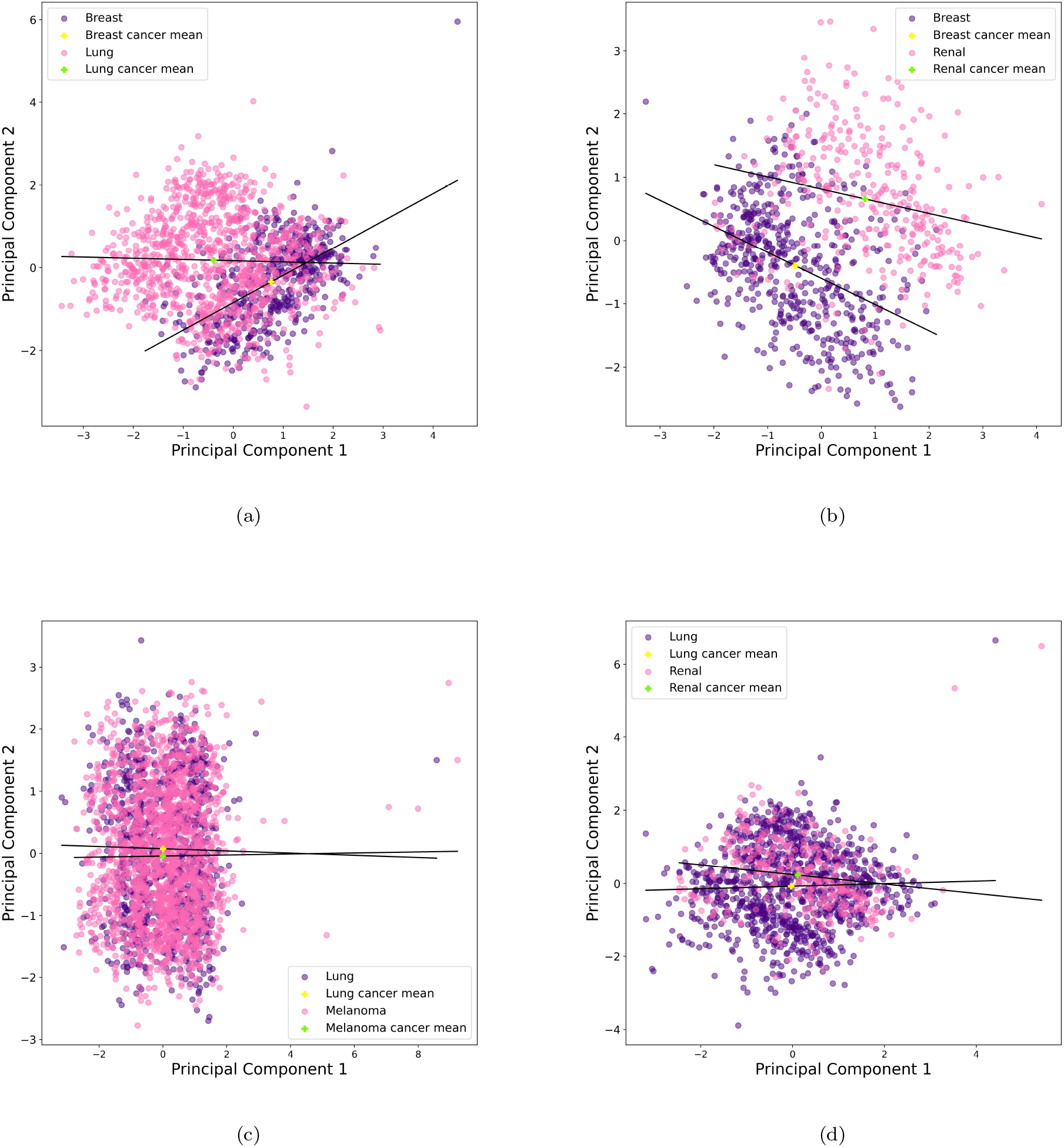
Scatter plot of pair cancer types onto (PC1, PC2) axes. The black line indicates the linear curve fit is not meant to imply that the data is spread linearly, but is useful to draw attention to differences in the two data sets being compared. Yellow and green crosses show the means. a, and b plots have the most distinct brain metastasis distributions (lung vs. breast cancers, and breast vs. renal cancers), c, and d have the most similar brain metastasis distributions (lung vs. melanoma cancers, and lung vs. renal cancers).

We now use three different Machine Learning, and Deep Learning algorithms: Random Forest model, Support Vector Machine (SVM) and TabNet to see how well each can distinguish between the BM spatial distributions associated with the five primary cancer types. See discussions of these algorithms in the Methods section.

The first important observation is shown in figure 4 where we plot the relative importance of the top 8 most important features from the data. PC1 is identified as the most important feature, followed by the Z coordinate, the Y coordinate, then PC2, PC3, followed by Age at treatment, X coordinate, and Target volume. Taken together, our conclusion is consistent with our previous observations, that (PC1, PC2) are a more efficient coordinate system to use than (Y,Z) given that the PC1 direction captures most of the spread in the (Y,Z) plane. In addition, Age at treatment seems to be a more important variable than Target volume in distinguishing spatial BM distributions. Table III summarizes key metrics (Accuracy, Precision, Recall, F1-score) associated with the Random Forest (RF) method, and three different Support Vector Machine (SVM) methods: SVM-linear; SVM-poly; and SVM-rbf as well as TabNet. With all metrics, the SVM-poly method performs best, scoring at 97% on the test data in each category. In Table III, Precision is the number of correctly identified members of a class divided by the number of times the model predicted that class; Recall is the number of members of a class that the classifier identified correctly divided by the total number of members in that class; and F1-score is a combination of Precision and Recall combined into one single metric. By contrast, SVM-linear performs the poorest in each category.

**FIG. 4.**
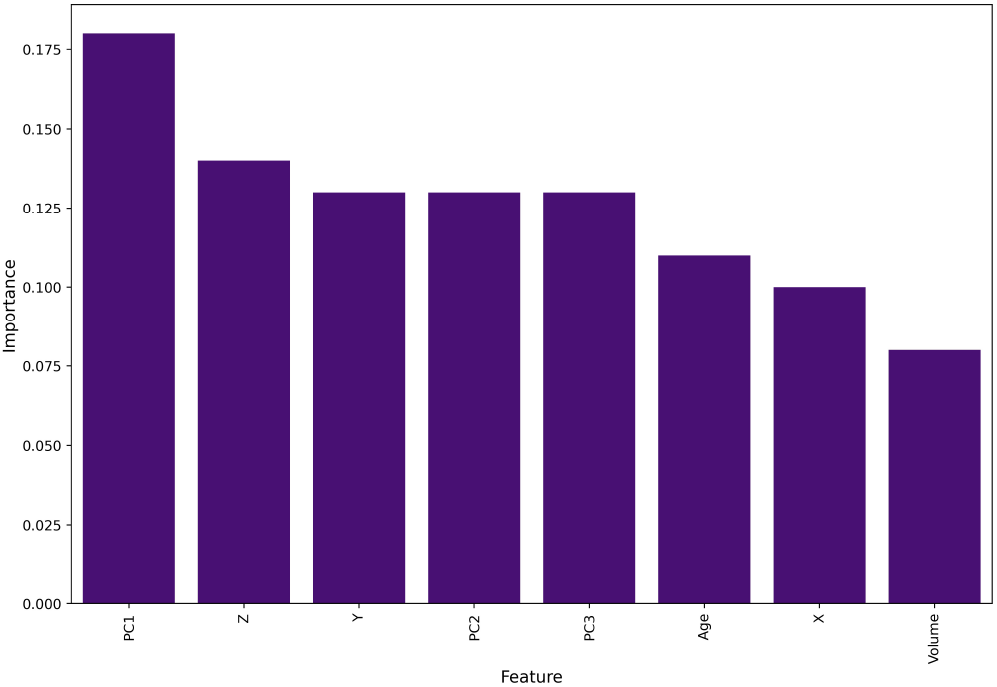
Bar plot showing the feature importance of the Random Forest model. (PC1,PC2,PC3) coordinate features are collectively more important than (X,Y,Z) coordinate features. Age at Treatment is a more important non-coordinate feature than Target volume.

## IV. DISCUSSION

The primary objective of this study was to analyze the spatial distribution of brain metastases (BMs) across several primary cancers to test whether machine learning models can discern differences among them. The results demonstrated that the first principal component (PC1) exhibits a significant alignment with the Y and Z axes, with minimal correlation observed with the X axis. Furthermore, our machine learning models achieved high accuracy in distinguishing spatial distributions of BMs for different primary cancers, with the Support Vector Machine (SVM) algorithm using a polynomial kernel achieving a 97% accuracy rate using all the standard metrics. In this discussion, we will interpret the principal component analysis, compare the primary cancer types, discuss the machine learning analysis, explore clinical implications and applications, and address limitations and future research directions.

### A. Interpretation of Principal Component Analysis

The principal component analysis (PCA) performed in this study aimed to reduce the dimensionality of the dataset in an optimal way [12], allowing for easier visualization and interpretation of the spatial distribution of BMs across the different primary cancer types. Preparing the data using PCA is key to achieving high metrics for the machine learning models. The importance of the first principal component (the axis of the largest spread of the data) in distinguishing the spatial distribution of BMs was demonstrated by its strong correlation with the Y and Z axes (figures 1(a), 1(b)), which represent the front-to-back and top-to-bottom dimensions of the brain, respectively. In contrast, the X axis, representing the side-to-side dimension, showed minimal correlation with PC1 (figure 1(c)) or utility as a distinguishing feature (figure 4) presumably due to the similarities in the left-right symmetriesin the distributions for all of the primary cancer types. By contrast, as shown in figure 4, PC1 was identified as the most important feature of the RF algorithm.

The use of the (PC1, PC2) plane as an optimal reduced dimension plane allowed us to best depict the differences in the spatial distributions of BMs for the five different primary cancer types (Breast, Lung, Melanoma, Renal, and Colon). The (PC1, PC2) plane not only captured the important differences in Cartesian BM locations but also proved to be more efficient than the (Y, Z) coordinate plane which is not surprising due to the fact that the principal coordinate system is the optimal data-based coordinate system that captures the primary directions of spread (standard deviations) in the data.

### B. Comparison of Primary Cancer Types

Our analysis revealed distinct differences in the spatial distributions of BMs for Breast vs. Lung and Breast vs. Renal cancer types of cancers [21]. The linear curve fits for each of these cancer types on the (PC1, PC2) plane (figures 3(a), 3(b)) showed differences in both their means and orientations. These findings suggest that the spatial distribution of BMs may be more specific to the primary cancer type. We have similarly demonstrated these types of differences in genetic subtypes of breast cancer [9], but further exploration is required to understand the cellular or genetic mechanisms which may explain these spatial differences as discussed, for example in [11].

In contrast, the spatial distributions of BMs for Lung vs. Melanoma and Lung vs. Renal cancer types were found to be most similar of the ones we compared, as evidenced by the similarity of their means and linear curve fits on the (PC1, PC2) plane (figures 3(c), 3(d)). These similarities suggest that BMs originating from these primary cancer types may share common mechanisms or pathways in their development and progression, although this remains to be fleshed out.

### C. Machine Learning Analysis: Distinguishing Primary Cancer Subtypes

The application of machine learning algorithms, namely Random Forest (RF), Support Vector Machine (SVM), and TabNet deep learning, allowed us to assess their ability to accurately distinguish between the subtypes of primary cancers based on the spatial distribution of brain metastases (BMs). The high accuracy achieved by these models in most cases not only suggests the presence of distinct differences in the spatial distribution of BMs across primary cancer types but also indicates that the translation of these distributions onto the first principal component (PC1) further enhances the differentiation capabilities as indicated by its standing as the most important feature in the RF algorithm. This observation implies that utilizing the PC1, which already highlights differences in spatial distribution, can be a robust approach for parsing out these distinctions among primary cancer subtypes and should be an important component in using ML methods on larger data sets.

The downstream effects of developing ML and DL models for BM subtyping could be multifold. For one, while patients with BM often have known primary cancer diagnoses, there are often instances where neurologic symptoms and brain MRI are the first scans which demonstrate tumor burden. A high-fidelity test could at the minimum, key in radiologists and oncologists to look out for a particular subtype. Second, by addressing phenotypic, tumoral behavior characteristics (e.g. where it metastasizes), and exploring molecular traits which have overlap irrespective of primary cancer subtype (i.e. where it came from), we may unlock new options for therapeutic targets that are shared between seemingly disparate cancer subtypes.

## V. CONCLUSIONS

For the purposes of distinguishing the spatial distribution of brain metastases associated with the five primary cancer types under study, we find that the optimal data-designed coordinates PC1 vs. PC2, as opposed to the Cartesian Y-Z coordinate plane offers the best reduced dimensional projection in which to highlight differences in the spread of the BM data. As a variable in our feature-based machine learning approaches, PC1 emerges as the single most important feature to distinguish the spatial patterns. Instead of the (X,Y,Z) features in our ML approaches, the best set of features to use are (PC1, PC2, PC3), with Age at Treatment being more important than Target volume, but less important than the PC variables. The SVM-poly ML method performs very well (97% on test data by all metrics) in distinguishing among the five cancer types based on their BM distributions. We believe with more data, and better optimization of the ML and DL pipelines, ML and DL methods offer a very promising approach towards discerning potentially subtle differences in BM distributions associated with primary tumor type.

## Data Availability

All data produced in the present study are available upon reasonable request to the authors

## ACKNOWLEDGMENTS

Partial funding through the USC Norris Comprehensive Cancer Center’s Multi-Level Cancer Risk Prediction Models pilot Project Award, ‘Molecular, Clinical and Neuro-imaging Determinants of Spatiotemporal Pathogenesis of Cancer-Specific Brain Metastases: Data Analysis and Longitudinal Modeling’ (12/01/2020-11/30/2021) is gratefully acknowledged.

## Notes

### Competing Interest Statement

The authors have declared no competing interest.

### Funding Statement

Partial funding through the USC Norris Comprehensive Cancer Centers Multi-Level Cancer Risk Prediction Models pilot Project Award, Molecular, Clinical and Neuro-imaging Determinants of Spatiotemporal Pathogenesis of Cancer-Specific Brain Metastases: Data Analysis and Longitudinal Modeling (12/01/2020-11/30/2021) is gratefully acknowledged.

### Author Declarations

USC Biomedical IRB waived ethical approval for this work.

